# PrecisionPro Fusion: Clinical Validation of an Automated MRI-CT Fusion System for Prostate Radiotherapy Planning

**DOI:** 10.1101/2025.07.17.25331682

**Authors:** Deondre Do, Christopher Conlin, Madison Baxter, John Christodouleas, Robert Dess, Irena Dragojevic, Mukesh Harisinghani, Sophia Kamran, Vitali Moiseenko, Himanshu Nagar, Nabih Nakrour, Lily Nguyen, Rhea Rupareliya, Steven Seyedin, Yuze Song, Anders M Dale, Tyler M Seibert

## Abstract

**Background:** Accurate image registration between magnetic resonance imaging (MRI) and computed tomography (CT) is required for precise radiation therapy of prostate cancer. Manual registration methods have been identified as a significant barrier to the implementation of advanced treatment techniques such as focal boost therapy.

**Purpose:** To evaluate the accuracy of PrecisionPro Fusion—an automated MRI-CT registration pipeline— compared to manual registration by experienced radiation oncologists.

**Materials and Methods:** We conducted a prospective, multi-institutional validation study involving six genitourinary radiation oncologists from three institutions who performed registrations on 20 patient cases. The study used a two-round design with a one-month washout period, where physicians conducted MRI-CT registrations with and without PrecisionPro Fusion. We compared PrecisionPro Fusion to practical accuracy limits of manual registration, defined by intra-physician variability (distance between a physician’s two MRI-CT registrations of the same patient case) and inter-physician variability (maximum distance between a physician’s registration and the physician consensus— average of all physicians’ registrations of that patient case). Physician participants reported on the PrecisionPro Fusion user experience using a System Usability Scale questionnaire.

**Results:** Intra-physician variability for manual subspecialist registrations was median 2.9 mm (IQR: 1.9, 5.4); inter-physician variability was median: 4.7 mm (4.3, 5.7). PrecisionPro Fusion registrations had median distance from the physician consensus of 1.3 mm (IQR: 0.9, 2.7). The system received high usability scores (median 81; IQR: 74, 88).

**Conclusion:** PrecisionPro Fusion provides prostate MRI-CT registration accuracy comparable to manual physician registration. Automated MRI-CT registration could enable faster delineation of structures visible on MRI, including the urethra and intraprostatic tumors.

## 2 Introduction

Maximizing radiotherapy efficacy while sparing surrounding healthy structures depends on accurate delineation of targets and normal organs [1-4]. Physicians generally delineate the prostate on CT scans used for dose calculation, but because the prostate is poorly visible on CT (due to limited soft tissue contrast), substantial errors are very common [5,6]. MRI offers a superior view of the internal anatomy and facilitates improved segmentation accuracy and consistency [7,8]. Additional benefits of MRI-based radiotherapy include ability to focally boost dose to the MRI-visible tumor (to improve treatment efficacy) [9,10] and to spare the urethra (to reduce genitourinary toxicity) [11]. To take advantage of the improved anatomic accuracy of MRI and CT-based electron density maps, it is typically necessary to register (fuse) MRI to CT. Specifically, the *prostate* must be aligned between the two types of images, which is made challenging by the very different appearance of the modalities, variable patient setup (diagnostic MRI is often performed with empty bladder on a curved table top, compared to full bladder and flat couch for radiation CT simulation), natural physiologic changes in internal anatomy (e.g., variable rectal gas), and lack of reliable anatomic soft tissue landmarks.

Radiation oncologists view MRI-CT registration as a major obstacle to wider adoption of focal tumor boosting, limiting implementation of a technique with level 1 evidence of improved clinical outcomes [12, 13]. Current registration methods are manual and time-intensive; they also require specialized expertise to locate the prostate, even on MRI [14]. Standard registration methods rely on bony landmarks that may not accurately reflect the position or orientation of the prostate, which can move between imaging sessions due to variations in bladder and rectal filling, patient positioning, or natural organ mobility, making bone-based registration inherently unreliable for precise prostate localization. Manual registration can take a physician as long as 10 to 30 minutes, depending on expertise, which also presents an efficiency barrier [15]. These limitations highlight the inadequacy of conventional registration approaches and underscore the need for multi-modal imaging strategies for organ-specific registration.

An automated MRI-CT registration workflow that performs comparably to expert radiation oncologists could accelerate adoption of MRI in routine prostate cancer treatment planning, facilitating accurate prostate targeting, urethra sparing, and tumor boosting. One classical technique is the use of pre-defined anatomical templates (atlases) to facilitate information transfer between the two modalities, including for multimodal registration [16-18]. An atlas-based registration approach could be augmented by recent advancements in artificial intelligence (AI), which can provide accurate automated prostate contours on MRI [19]. We therefore developed PrecisionPro Fusion, an automated, *prostate-specific* MRI-CT registration pipeline. This system combines an atlas-based MRI-CT registration approach with a deep-learning-based prostate auto-segmentation in a multi-stage framework to combine the benefits of both techniques and allow for a seamless automated MRI-CT image registration workflow.

In this study, we evaluated the performance of PrecisionPro Fusion in a multi-institutional study, comparing PrecisionPro Fusion’s fully automated registrations to manual registrations by radiation oncologists who subspecialize in treatment of prostate cancer. We hypothesized that PrecisionPro Fusion would yield registrations accurate to within the practical limits of human accuracy (as defined by intra-and inter-physician registration variability).Accurate automated registration could improve efficiency everywhere and increase registration confidence in centers less experienced in MRI-based prostate cancer treatment.

## 3 Materials and Methods

Development and validation of PrecisionPro Fusion included imaging data from 3 institutions (Figure 1A).

**Figure 1.**
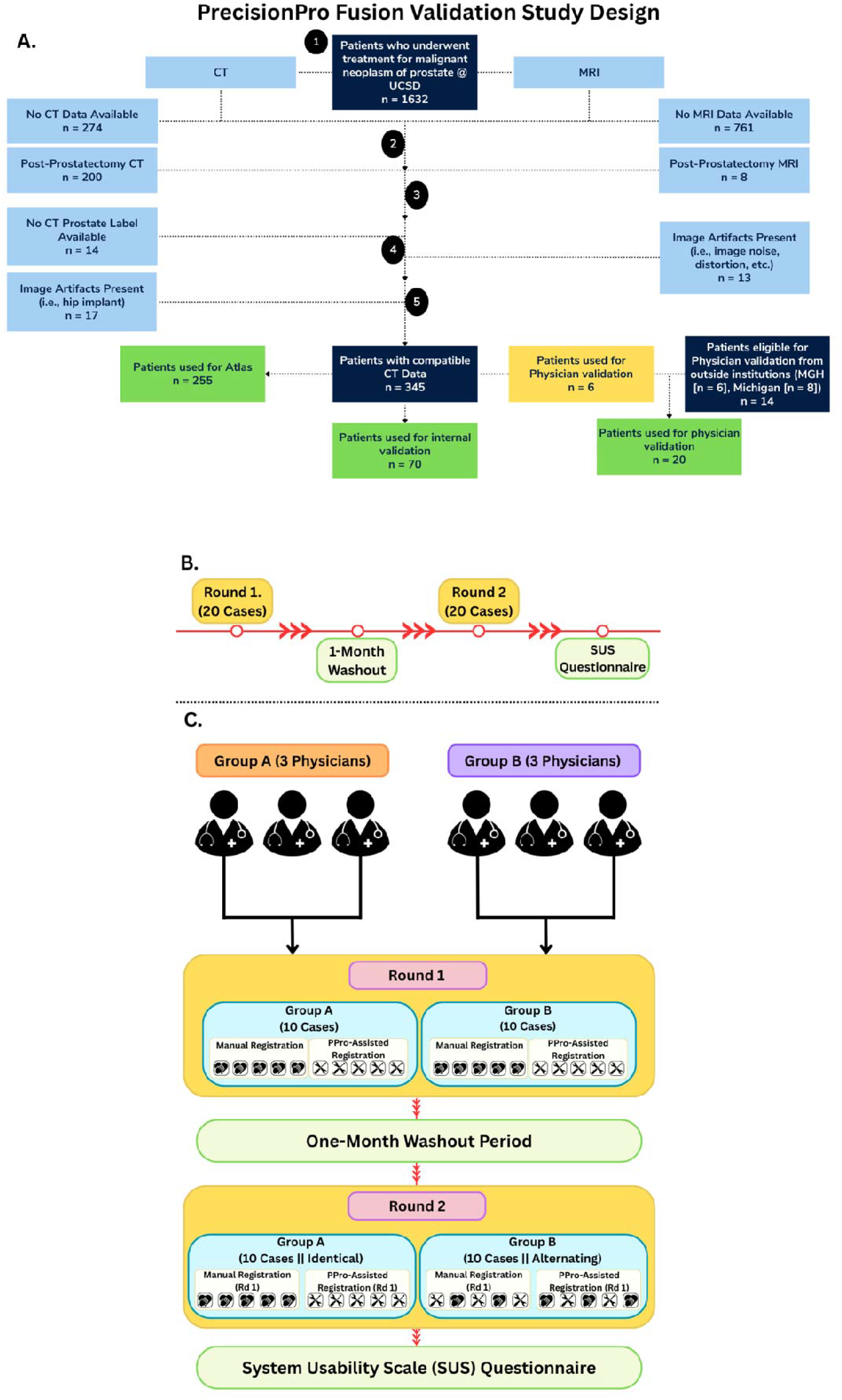
Multi-institutional patient cohort flow diagrams for PrecisionPro Fusion Validation Study. (A) Patient flow diagram showing the distribution of 325 patients included in this study: 255 patients from Institution 1 [UCSD] were utilized for atlas development, with an additional 70 patients from the same institution serving for internal validation. Prospective physician validation involved 20 patients distributed across three institutions with 14 from outside Institution 1 (6 from Institution 1 not used for the atlas or internal validation, 6 from Institution 2 [MGH], and 8 from Institution 3 [Michigan]).(B) Timeline overview of the study, consisting of two rounds of registration done by six subspecialist genitourinary radiation oncologists (10 cases per physician per round), separated by a 1-month washout period. (C) The validation study workflow for two physician groups (n=3 each). In Round 1, both groups performed manual registrations and PrecisionPro Fusion-assisted registrations on 10 cases each (5 manual and 5 assisted). Following a one-month washout period, Round 2 was conducted with Group A receiving identical cases (with identical registration methodology from Round 1), while Group B received identical cases but alternating registration methodology. This crossover design with washout period minimizes memory bias and allows assessment of both intra-physician and inter-physician variability. The study concluded with a System Usability Scale questionnaire evaluation to measure the system’s perceived usability by clinical experts.

### 3.1 Atlas Development

We created MRI and CT atlases using data from patients treated at Institution 1 (University of California, San Diego [UCSD]) between 2017 and 2023, excluding those with hip implants, significant image artifacts, or were treated without MRI. Approximately 20% of the cases were held out from atlas development to use for internal validation. Radiation planning CT scans had been annotated with physician-approved prostate contours. We applied a commercially available automated prostate segmentation tool (OnQ Prostate, Cortechs.ai, San Diego, CA) to *T*_*2*_-weighted MRI [20]. The atlas creation process began by calculating the centroid of each patient’s prostate segmentation, providing a consistent reference point. We aligned each patient’s MR images and segmentations by registering their prostate centroids to the centroid of a reference image, created as an empty volume with standardized dimensions in MATLAB (400×400×100mm). As the atlases use the same reference image volume, they are inherently registered at their centroid location (200×200×50mm). Once all patient images were registered to this common image space, we created the atlas by averaging the voxel intensities of the registered MR images and their respective prostate segmentations after translation and scaling.

We repeated this process for CT images and segmentations. The atlases are probabilistic maps representing typical prostate appearance and position across both imaging modalities.

### 3.2 PrecisionPro Fusion Registration Framework (Supplementary material 1A)

When registering a new patient’s MRI and CT images, PrecisionPro Fusion employs a two-step approach:

1. **MRI Registration**: First, we generate a prostate segmentation on the patient’s axial T2-weighted MRI using an in-house deep learning tool previously shown to have high accuracy [8]. a. We calculated the centroid of this prostate segmentation and determined the translation needed to align it with our MRI atlas’ prostate centroid as described above.
2. **CT Registration**:

a. First, we detected an anatomical landmark in the pelvic region to estimate the prostate location on the patients’ CT image and create a rough rigid registration with our CT atlas.
b. Second, we performed a multi-stage optimization process that aligns the patient’s CT with our CT atlas using a cost function with a weighted combination of intensity-based and statistical metrics.

Because our MRI and CT atlases are inherently aligned, registering each modality’s images to the respective atlas created a bidirectional mapping between the patient’s MRI and CT at the prostate’s location. Additional technical details are described in the Supplementary Material.

### 3.3 Internal Validation

We conducted an initial comparison by applying automatic registration tools using the vendor-supplied mutual information-based rigid registration algorithms in MIM and Eclipse (Varian Medical Systems, Palo Alto, CA) and comparing these registration outputs to those from PrecisionPro Fusion. The automated (bone-based) vendor algorithms consistently failed to produce a valid registration due to the significant differences in the field-of-view between the diagnostic MRI and planning CT images (Supplementary Figure 1)

We further performed an internal validation using an independent cohort of patients from Institution 1 who were not included in the atlas development dataset. PrecisionPro Fusion was applied to each patient case in this dataset and then visually evaluated for registration accuracy, with the aid of automated prostate segmentation on MRI and the physician-approved manual prostate contours on CT.

### 3.4 Prospective Validation

We conducted a prospective, multi-institutional validation of PrecisionPro Fusion by comparing its automated registration results to manual registration performed by physicians (Figure 1B, 1C). Six subspecialist genitourinary radiation oncologists from six US academic medical centers participated in two rounds each, separated by a one-month washout period.

In Round 1, physicians performed MRI-CT registration for 10 patient cases: half without PrecisionPro Fusion and half using the PrecisionPro Fusion registration as an initial starting point, with review and adjustment if needed. There were 20 patient cases in all, with pairs of physicians randomly assigned 10 of the cases so that there were always at least two physician registrations of each case. After the washout period, Round 2 involved the same task: registration of the same 10 cases as they saw before, though in randomized order. Additionally, the physician participants were assigned to one of two groups: Group A received identical cases in both rounds, while Group B received half identical cases and half cases switched from manual to PrecisionPro Fusion between rounds to allow for within-subject comparisons.

Registration tasks were performed using MIM (MIM Software, Beachwood, OH). All physician participants received training for how to use MIM to perform and save their registrations through brief self-conducted tutorials (text and video), with personal Q&A available if needed. Physicians were also asked to report their user experience with the PrecisionPro Fusion outputs using the standardized System Usability Scale (SUS) at the end of the study.

### 3.5 Measurement of Registration Accuracy

Given there is no clear ground truth for prostate registration accuracy, we adopted a pragmatic approach (Figure 2). Registrations judged by subspecialist physicians to be accurate enough for clinical use are considered to have cleared the bar of clinical utility. We found the centroid of all physician manual MRI-to-CT registration matrices for each patient case, yielding the average answer across physician participants. We then measured the Euclidean distance (in mm) between the prostate center of mass (based on auto segmentation on MRI) for each physician’s registration and the average position across physicians. Likewise, we measured the distance between the PrecisionPro Fusion registration (without manual adjustment) for each patient case with the physician consensus/average. The PrecisionPro Fusion distance was compared to intra-physician variability (distance between the same physician’s Round 1 and Round 2 registrations) and inter-physician variability (maximum distance between a physician’s registration and the physician consensus). Registrations were rigid (no shrinking/expanding), with left-right, anterior-posterior, and superior-inferior directions evaluated. Rotation and deformation were not considered/applied.

**Figure 2.**
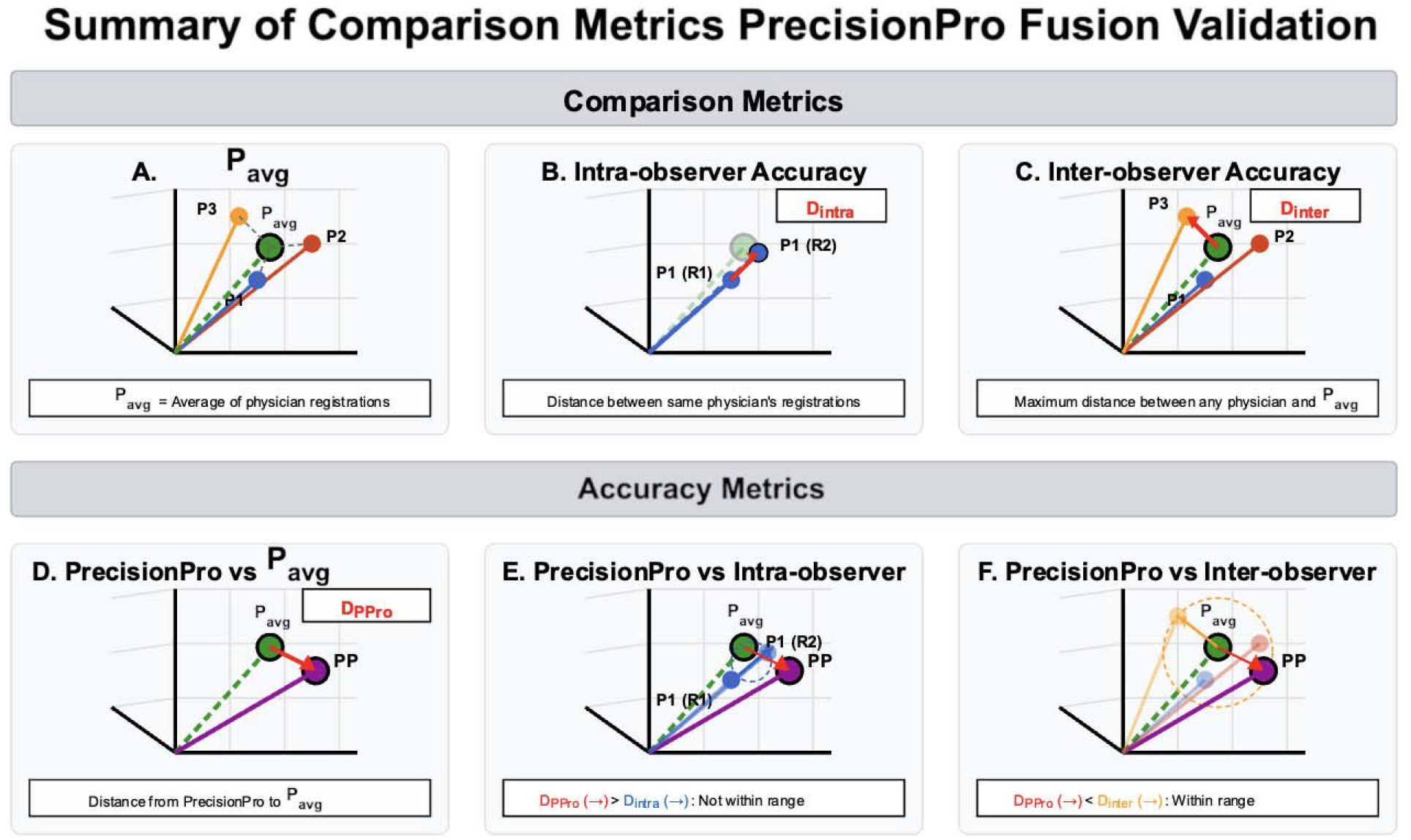
PrecisionPro Fusion Validation. Quantitative Comparison Metrics Top row (A-C) demonstrates the metrics used for comparison: (A) The consensus (P_avg_) determined as the average of three physician registrations (P1, P2, P3) for each individual patient case; (B) Physician intra-observer variability (D_intra_) is measured as the distance between a physician’s repeated registrations between Round 1 and Round 2 (i.e., P1-R1,P1-R2); and (C) Physician inter-observer variability (D_inter_) is measured as the maximum distance between any physician’s registration and P_avg_.**Bottom row (D-F)** shows how we evaluated clinical acceptability of PrecisionPro Fusion’s registrations for each case: (D) PrecisionPro Fusion’s (PP) registration compared directly to P_avg_, with the distance between the two points defined as D_PPro_; (E) Comparison of D_PPro_ to D_intra_, showing that PrecisionPro Fusion’s registration accuracy exceeds the range of intra-physician variability, we would consider this registration a failure to meet clinical acceptability; and (F) Comparison of D_PPro_ to D_inter_, demonstrating that PrecisionPro Fusion’s accuracy falls within the acceptable range of inter-physician variability and is clinically acceptable for use in treatment planning.

### 3.6 Statistical Analysis

Our analysis included a quantitative comparison of PrecisionPro Fusion’s registration versus the range of intra-/inter-physician variability to determine the accuracy of our registration pipeline. Distance from the physician consensus was also calculated to quantify accuracy as explained above. We further analyzed how intra-/inter-physician variability changed when using PrecisionPro Fusion and analyzed the directional variability of physician adjustments to the initial PrecisionPro Fusion registrations across the three cardinal coordinate axes. We calculated median values with interquartile ranges (IQR) as our primary statistical descriptors. PrecisionPro Fusion registration was considered successful if distance from the physician consensus fell within the observed ranges of manual intra-/inter-physician variability. A SUS assessment was conducted at the end of the study to evaluate user experience, with scores above 68 considered above average and indicative of acceptable clinical usability [21].

## 4 Results

Data from 325 patients were used in this study, split into three cohorts. 255 patients from Institution 1 were used to develop the MRI and CT atlases, and 70 patients from Institution 1 were used for internal validation and testing of PrecisionPro Fusion. Data from 20 additional patients selected from 3 participating institutions—6 from Institution 1, 6 from Institution 2 (Massachusetts General Hospital at Harvard [MGH]), and 8 from Institution 3 (University of Michigan [Michigan])—were reserved for the prospective physician validation to evaluate performance between different scanners and protocols. Final validation was performed in MRI data from 5 different scanners and 2 vendors and CT data from 3 different scanners and 3 vendors (Supplementary Table 1).

**Table 1.**
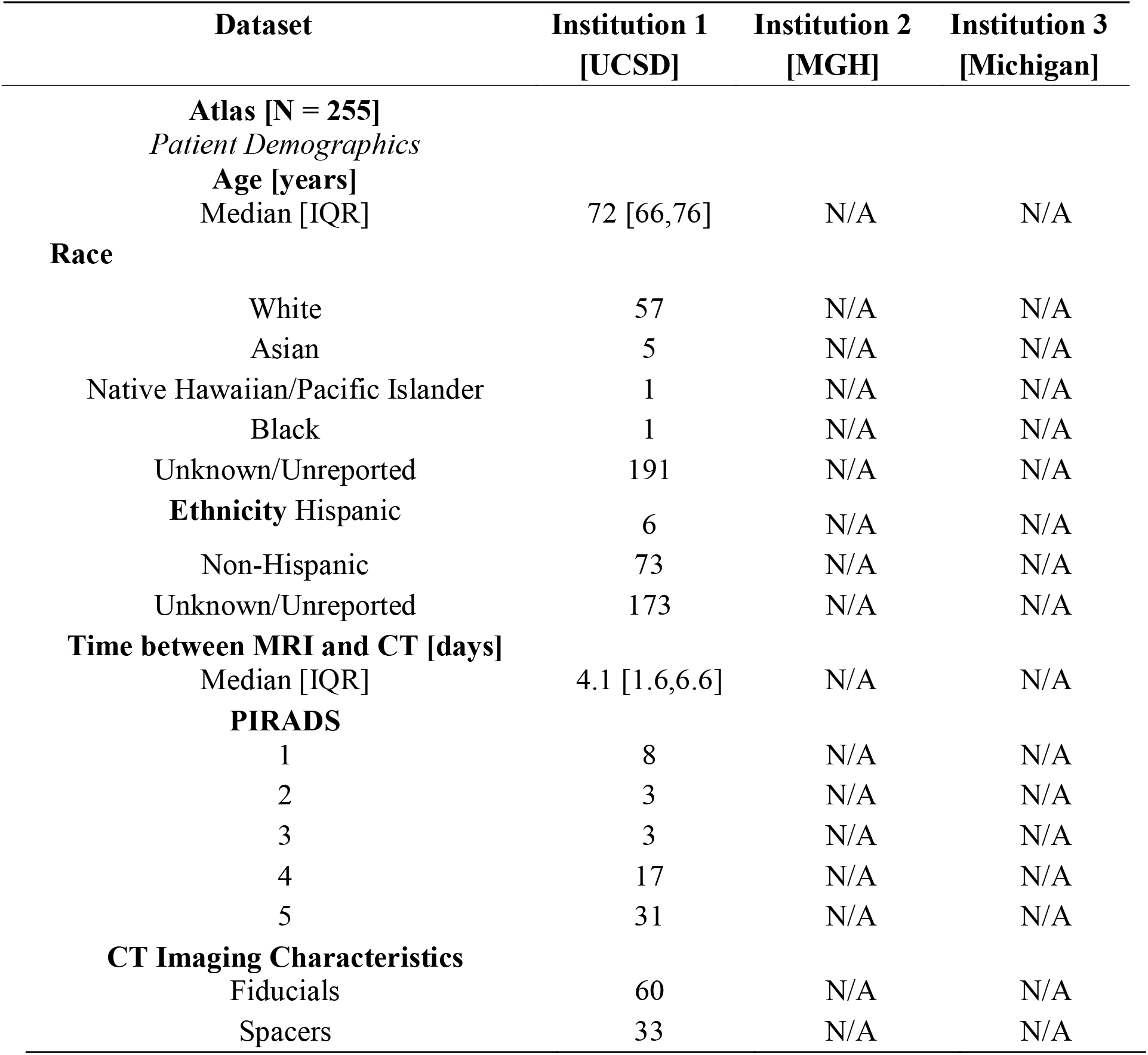
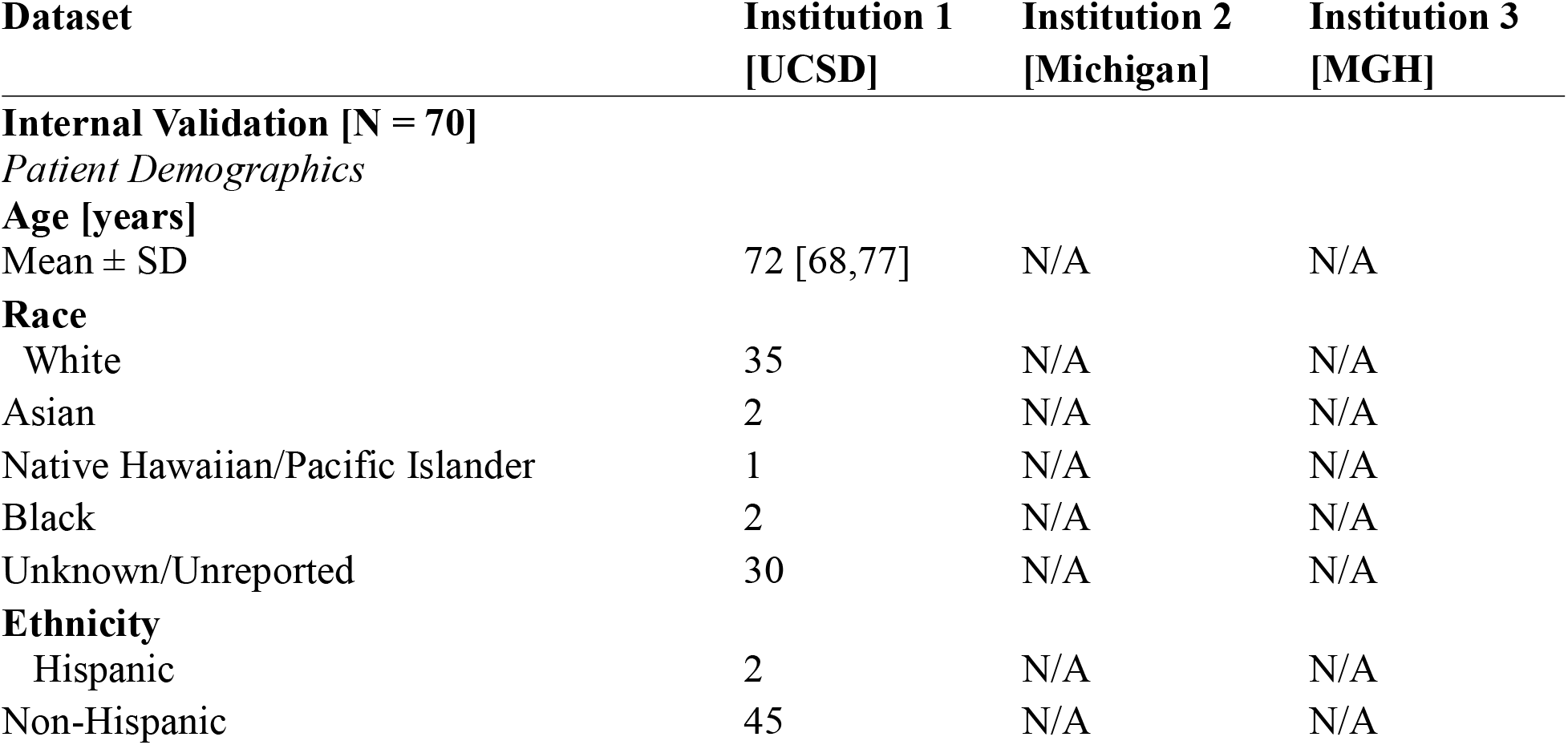
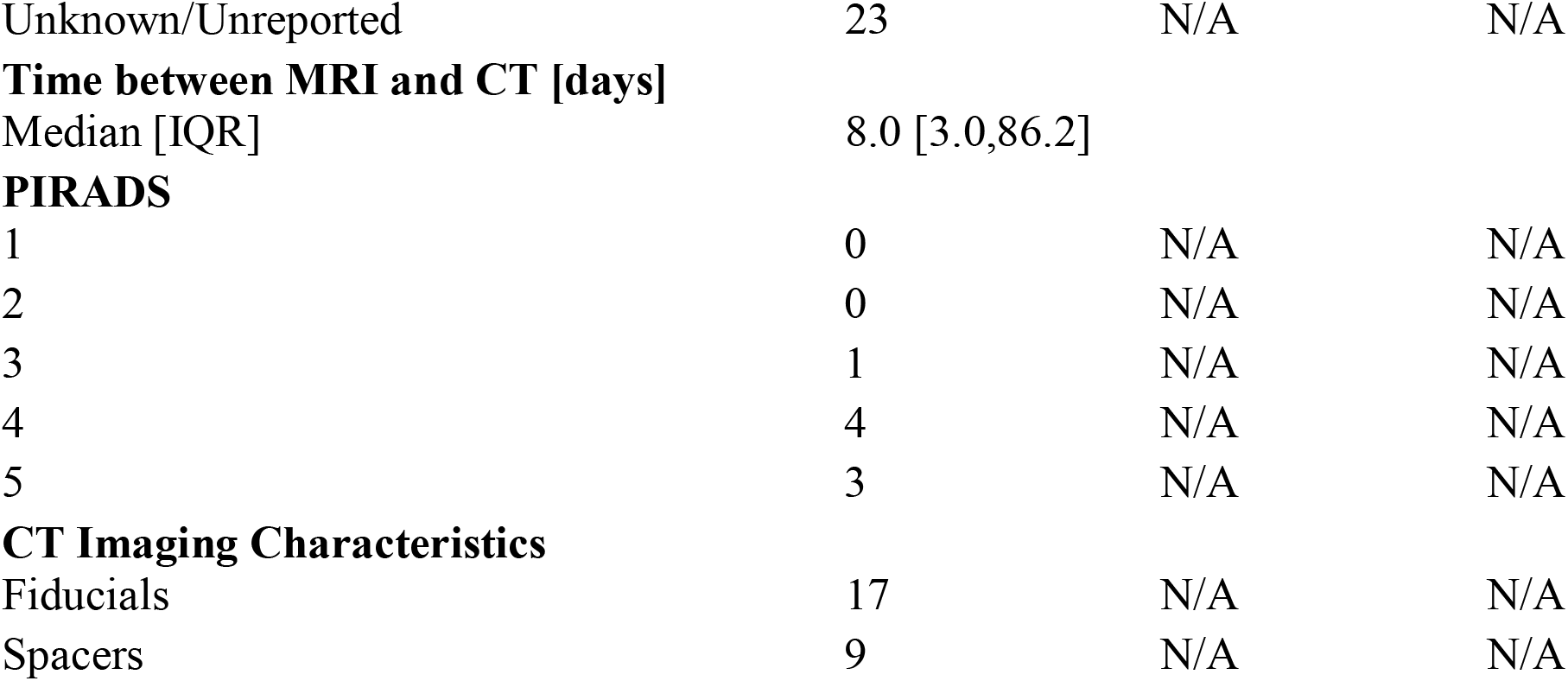
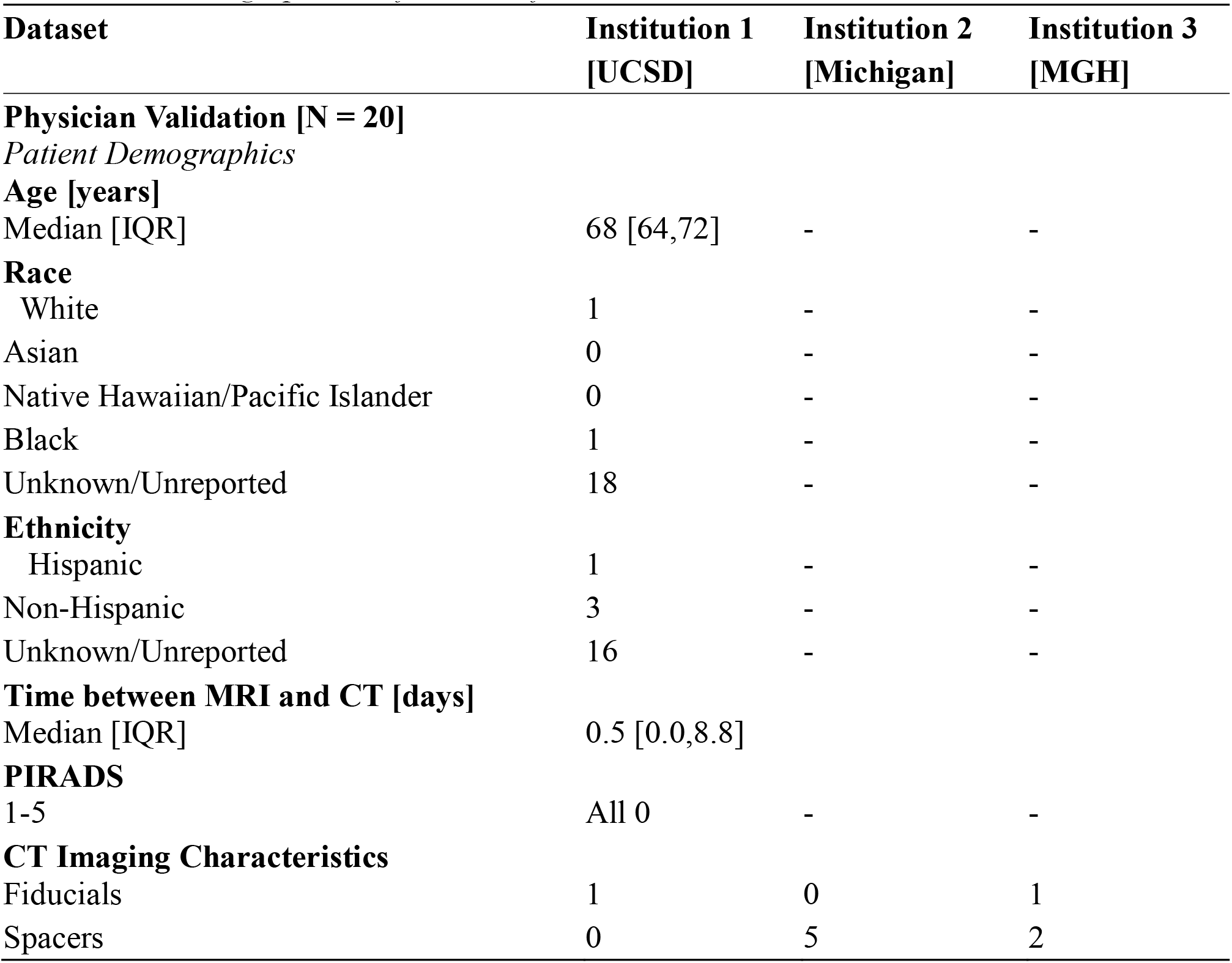
Patient Demographics and Imaging Characteristics Across Institutions. *\*Only UCSD patients were used for the atlas development and internal validation of PrecisionPro Fusion* *\*Demographical information from Institutions 2 and 3 were unavailable*

### 4.1 Primary Outcomes: Registration Accuracy Comparison

Physician manual registration performance demonstrated an inter-physician variability with a median of 4.7 mm (IQR: 4.3, 5.7 mm) and round-to-round intra-physician variability with a median of 2.9 mm (IQR: 1.9, 5.4 mm) (Figure 3). Thus, the practical limit of measurable accuracy is 3-5 mm.

**Figure 3.**
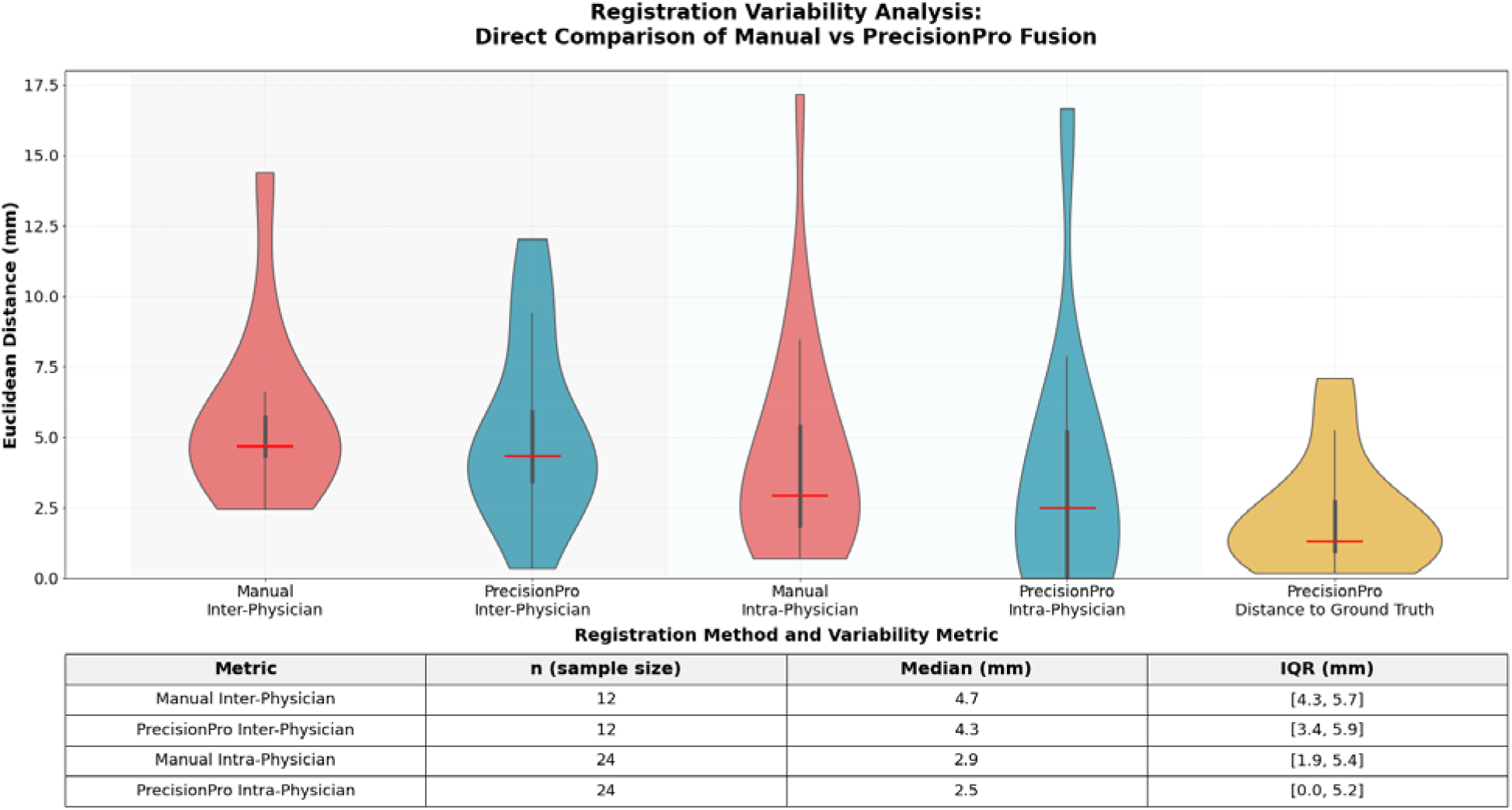
Comparative analysis of manual inter-physician and intra-physician MRI-CT registration variability vs PrecisionPro Fusion-assisted alignment. Violin plots illustrating the distribution of registration variability among different (inter-), and the same (intra-) physicians of manual (red) compared to PrecisionPro Fusion-assisted (blue) MRI-CT registration. Manual inter-physician variability had a median deviation of 4.7 mm (IQR: 4.3,5.7 mm) and manual intra-physician variability demonstrated a median deviation of 2.9 mm (IQR: 1.9,5.4 mm), suggesting a range between 3 and 5mm for physicians conducting MRI-CT registration is reasonable as a practical limit of measurable registration accuracy. Inter-/Intra-physician registration variability (blue) using the PrecisionPro Fusion platform achieved a median deviation of 4.3 mm (IQR: 3.4,5.9 mm) and a median deviation of 2.5 mm (IQR: 0,5.2 mm) respectively, indicating a potentially enhanced reproducibility for repeated registrations by the same physician when using PrecisonPro Fusion compared to without it. PrecisionPro Fusion demonstrated high accuracy (yellow) compared to the physician consensus (average of all physicians’ registrations) with a median distance of 1.3 mm (IQR: 0.9,2.7 mm) from the physician consensus.

PrecisionPro Fusion registrations gave a median distance from the physician consensus of 1.3 mm (IQR: 0.9, 2.7 mm). In 10 of 12 patient cases, PrecisionPro Fusion registrations were closer to the physician consensus than the observed intra-physician variability for the patient case (Supplementary Figure 2). Inter-physician variability using the PrecisionPro Fusion workflow was median 4.3 mm (IQR: 3.4, 5.9 mm), while round-to-round intra-physician variability was median 2.5 mm (IQR: 0.0, 5.2 mm).

When physicians were provided PrecisionPro Fusion results as a starting point, they manually adjusted the registration position a median 3.0 mm (IQR: 0.0, 5.0), typically within the practical limit of measurable accuracy (Figure 4). There was a noticeably higher adjustment magnitude to PrecisionPro Fusion registrations specifically in the superior-inferior direction but otherwise no additional evidence for directional bias was found (Supplementary Figure 3).

**Figure 4.**
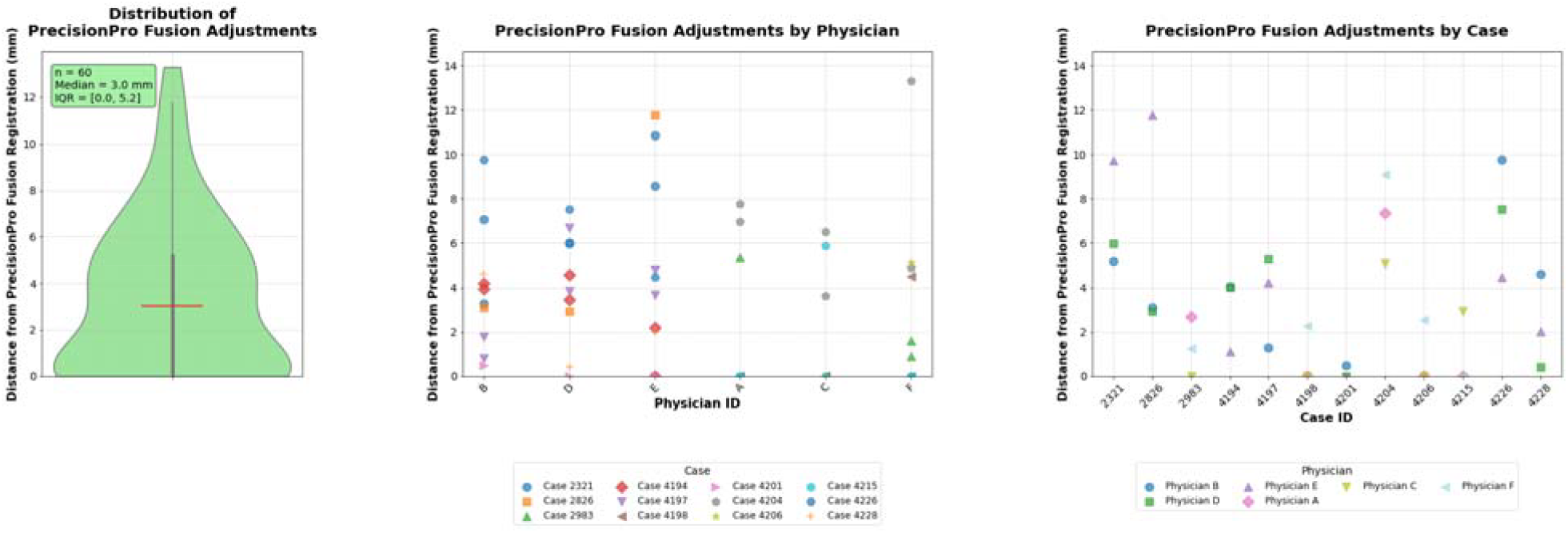
Quantitative assessment of physician adjustments to PrecisionPro Fusion registrations. The violin plot (left) shows the distribution of physician modifications to PrecisionPro Fusion, demonstrating a median displacement of 3.0 mm (IQR: 0.0,5.0 mm) from the initial registration position. Dot plots (middle and right) illustrate the magnitude of physician adjustments to PrecisionPro Fusion registrations, stratified by individual cases (left) and individual physicians (right). Notably, there are a few case-specific registration challenges (i.e., Case ID 2321) and physician-specific adjustment patterns (i.e., smaller adjustments by Physician C)

### 4.2 SUS Assessment

The SUS assessment yielded a median score of 81 (IQR: 74, 88).

## 5 Discussion

PrecisionPro Fusion achieved fully automated registration accuracy comparable to manual registration performed by experienced radiation oncologists who subspecialize in prostate cancer radiation therapy. The distance between PrecisionPro Fusion results and the physician consensus was typically smaller than intra-physician or inter-physician variability, and any adjustments to the initial PrecisionPro Fusion result was within the practical limits of measurable registration accuracy. Physician users rated PrecisionPro Fusion with high satisfaction.

A major challenge for analysis of MRI-CT registration of the prostate is the lack of an objective ground truth, especially on CT. We addressed this limitation by defining clinically acceptable results as registrations that fall within the inter-physician and intra-physician variability for experienced subspecialists. If the automated registration tool can achieve agreement with physician consensus that is within the round-to-round or physician-to-physician variability, we can conclude that the tool gives results that are as accurate as can be practically measured. Moreover, we note that physiological changes between MRI and CT scan events can make a perfect registration impossible. Fortunately, perfection is not necessary for clinical utility.

Rather, MRI-CT registration must be accurate enough for the physician to confidently use the information on MRI to inform treatment planning. All the physician participants in this study routinely use MRI to inform treatment planning for prostate radiation therapy, including to delineate the urethra, to improve contours of the prostate boundary, and to focally boost radiation dose to visible prostate tumors. PrecisionPro Fusion gives an automated starting point that is very close to the final subspecialist physician consensus, suggesting this tool could be useful in encouraging implementation of MRI-guided prostate radiation therapy.

Another approach would be the use of fiducial markers as a ground truth for registration accuracy [22]. Some patients in our study did have fiducials placed, but they were carbon markers that are highly visible on CT but inconsistently visible on MRI, making them unhelpful for the present analysis. Fiducial markers may move within the prostate, which could cause issues with registration, but they should generally improve registration accuracy if visible on both MRI and CT [23-25]. Not all centers place these routinely, though, due to lack of availability of an experienced physician to place them, cost, patient comfort, and risks of the invasive procedure [23].

A notable finding of this study was the manual inter-and intra-physician variability in manual registration, consistent with a previous study that highlighted significant variability in manual contouring and registration for prostate cancer [26]. This variability has clinical implications for advanced techniques like focal boosting and urethral sparing. The margins required to account for this user-dependent variability limits the benefits of these high-precision techniques, showing the clinical need for a robust registration tool that can produce consistent results, thereby reducing uncertainty and potentially allowing for smaller treatment margins.

PrecisionPro Fusion could help standardize and automate MRI-CT registration across institutions, addressing one of the major barriers to implementing advanced techniques like focal boost delivery [9]. This could be particularly useful in practice settings where registrations are typically performed first by dosimetrists or physicists who may not have previous training in prostate-specific MRI-CT registration. Physician supervision and adjustment should always be performed, but a decent initial starting point can be important for efficiency. Potential workflow efficiency gains offered by automated registration are particularly relevant in the context of increasing patient volumes and resource constraints in radiation oncology centers. Automated MRI-CT registration could be combined with automated segmentation of the prostate and automated treatment planning with focal radiation boost, both of which can perform as well as experienced humans [8,21,27].

Limitations of this study include a single institution for development of the atlas. PrecisionPro Fusion nonetheless performed well in multi-institution data. Logistics and physician reviewer time placed a limit on the feasible sample size for a prospective evaluation like this one. Still, we have enough observations to show that PrecisionPro Fusion registrations are within the practical limit of measurable accuracy, and the automated pipeline has potential to save physicians a large amount of time compared to manual registration. Indeed, it is unclear what a statistically significant improvement in accuracy could be when the results are already within the limits of measurable accuracy. A more relevant point is that a larger sample size would allow evaluation of a broader range of patient anatomy and imaging center variation to further inform generalizability of the results.

We also acknowledge that PrecisionPro Fusion does not account for potential prostate deformation between the MRI and CT scans. While deformable image registration theoretically represents a more sophisticated approach to address this issue, its application is complicated by the poor soft tissue contrast of planning CT making it difficult to find reliable corresponding features within the prostate, especially at the boundaries of the gland [28]. Prostate deformations and delineation uncertainties are therefore difficult to distinguish accurately, so a robust rigid registration of the whole prostate gland remains a common, pragmatic, and clinically relevant approach [15,29].

We note that scans with imaging artifacts (e.g., from hip implants or severe rectal gas effects on MRI) were excluded from this analysis. We are planning a larger, prospective implementation study where PrecisionPro Fusion and other tools to facilitate focal boost are evaluated for real-world utility in clinical workflows [8,21,30,31].

## 6 Conclusion

PrecisionPro Fusion achieves registration accuracy comparable to manual registration by experienced subspecialist radiation oncologists. The high system usability scores suggest potential for successful clinical adoption. These findings support the integration of automated registration in enhancing radiation therapy planning for prostate cancer and potentially facilitating the adoption of urethra avoidance and focal boosting of prostate tumors visible on MRI.

## Data Availability

All data in the present study are available upon reasonable request to the authors

