## Supplementary Material for "PrecisionPro Fusion: Clinical Validation of an Automated MRI-CT Fusion System for Prostate Radiotherapy Planning"

### 1A. PrecisionPro Fusion MRI-CT Registration Pipeline Methodology

This methodology describes a two-phase approach for automated MRI-CT registration of the prostate. First, we create probabilistic atlases from a training cohort that capture the statistical relationships between CT and T2-weighted MRI intensities, specifically in the prostate. Then, we use these atlases to register new patients' MRI and CT scans by leveraging both geometric alignment and intensity correspondence to these atlases.

### 1.1 Phase 1: Atlas Creation

The probabilistic atlases were constructed from a training cohort of 255 patients to establish a common reference space and statistical intensity relationships between imaging modalities.

#### 1.1.1 Data Preprocessing

For each training patient (i), CT and T2-weighted MRI volumes with corresponding manual prostate segmentations data underwent pre-processing before atlas creation.

**CT Preprocessing:** Raw CT volumes (I_CT_) in Hounsfield Units were standardized by scaling voxel intensities (v) of each training patient to a predefined range [b_min_, b_max_] (-150,200 HU):

I'_CT,i_(v) = min(b_max_, max(b_min_, I_CT,i_(v)))

**T2 MRI Preprocessing:** T2-weighted volumes (I_T2_) underwent an N3 bias field correction using an in-house algorithm (I'_T2_), followed by intensity normalization. The normalization scaled each volume so the median intensity within the prostate segmentation on MRI (S_T2_) became a constant value k_norm_ (set to 200):

I''_T2,i_(v) = I'_T2,i_(v) × (k_norm_ / median({I'_T2,i_(v') | v' ∈ S_T2,i_}))

This preprocessing ensures that intensity variations across patients reflect anatomical differences rather than acquisition-related factors.

#### 1.1.2 Geometric Alignment to Atlas Space

Training patient images were aligned to a common reference space through a two-stage geometric transformation designed to normalize both position and size variations.

**Stage 1 - Translational Alignment:** Each patient's prostate was centered by calculating the geometric centroid of the prostate segmentation:

C_i_ = (1 / |S_i_|) × Σ v for all voxels v ∈ S_i_

A translation transformation T_trans,i_ was applied to move each patient's centroid to the atlas reference center, ensuring consistent positioning across the cohort.

The complete transformation for each patient combined both alignment stages, including both scaling and translation.

T_final,i_ = T_scale,i_ ⋅ T_trans,i_

**Stage 2 - Size Normalization:** After centering, we computed a mean prostate shape from all aligned T2 segmentations:

S_mean_ = (1/n) × Σ S_centered,T2,i_

Individual scaling factors were then calculated by comparing each patient's prostate dimensions (w = width, h = height, d = depth) to this reference shape. Anisotropic scaling accommodated natural variations in prostate geometry:

s_x,i_ = w_ref_ / w_i_

s_y,i_ = h_ref_ / h_i_
s_z,i_ = d_ref_ / d_i_

Applying this transformation for each patient creates the final, centered volumes for atlas generation in CT image space:

I_CT,final,i_ = I'_CT,i_ x T_final,i_ x s_x,y,z,i_

I_T2,final,i_ = I''_T2,i_ x T_final,i_ x s_x,y,z,i_

S_final,i_ = S_i_ x T_final,i_ x s_x,y,z,i_

#### 1.1.3 Atlas Generation

The final probabilistic atlases (CT, T2 and accompanying segmentations) were created by averaging the aligned training data, capturing both mean intensities and statistical relationships:

A_CT_ = (1/n) × Σ I_CT,final,i_

A_T2_ = (1/n) × Σ I_T2,final,i_

A_S_ = (1/n) × Σ S_final,i_

Additionally, intensity standard deviation maps for each atlas and joint probability distributions P(I_CT_, I_T2_) between each atlas were computed to characterize the statistical relationships between modalities within the training cohort.

### 1.2 Phase 2: Patient MRI-CT Registration Using the Atlas

Once the probabilistic atlases are established, new patients can be registered by finding the optimal alignment between their scans and the atlas space. This multi-stage process leverages both geometric constraints and statistical intensity relationships to simultaneously register a patient’s MRI to CT and CT to atlas space to achieve a robust MRI-CT registration.

#### 1.2.1 Initialization and Data Preparation

**Atlas Component Loading:** The pre-computed probabilistic atlas components were loaded, including the mean intensity atlases (A_CT_, A_T2_), intensity standard deviation atlas (σ_CT_) and probabilistic T2 segmentation atlas (A_S_).

**Patient Data Processing:** Patient CT and T2-weighted MRI volumes underwent identical preprocessing to the atlas cohort: CT volumes were intensity-windowed and T2 volumes received N3 bias field correction and intensity normalization.

**Automated Segmentation:** Prostate segmentation masks were generated from the patient's T2-weighted MRI using an in-house automated deep learning tool, providing anatomical constraints for the registration process.

#### 1.2.2 Initial Affine Transformation Estimation

Both imaging modalities received initial transformations to bring them into approximate alignment with the atlas space before final optimization.

**T2 Volume Alignment:** The T2 volume alignment combined translation and anisotropic scaling during registration to atlas space. The geometric centroid of the patient's prostate segmentation was calculated and aligned with the atlas volume center via translation matrix T_trans_. Anisotropic scaling factors were derived by comparing a bounding box around the patient's prostate to the atlas prostate’s reference dimensions, forming a scaling matrix T_scale_. The complete initial T2 transformation was:

T_T2-initial_ = T_scale_ ⋅ T_T2-trans_

**CT Volume Initialization:** The patient's CT volume received initial scaling using the same T_scale_ matrix derived from the T2 prostate geometry, plus a small heuristic z-axis adjustment. This scaling ensured geometric consistency between modalities while preparing the CT for intensity-based optimization.

#### 1.2.3 Anatomical Landmark Detection

To obtain a robust starting point for the main optimization and avoid a computationally expensive search over the entire volume, a 3D coordinate is identified by automatically detecting prominent anatomical features in the patient's CT data. This process provides an initial estimate (x̂₀) to initiate the subsequent grid search. The procedure sequentially identifies the patient's central sagittal slice (y-coordinate), then the anatomical midline (x-coordinate), and finally a superior-inferior position (z-coordinate) at the prostate’s approximate location.

**2.3.1. Midsagittal Plane Identification**

The first step is to automatically identify the patient's central sagittal slice by finding the plane with the highest anatomical voxel heterogeneity.

1. **Variance Profile Calculation:** For each sagittal slice, s, a score, **V**_sag_(s), is computed. This score is the average of the intensity variances calculated for each vertical column of voxels (c) within that slice.

**V**_sag_(s) = (1/N) × Σ var(CT(:, s, c))

where N is the number of columns in the slice.

The central sagittal slice, **S**_mid_, is identified as the slice that yields the maximum variance. This slice corresponds to the y-coordinate, c_2_, for the initial estimate.

Additionally, a central slab of the body (**c**₂min, **c**₂max) is defined as the region where the variance is above 30% of its maximum value to determine the remaining coordinate estimates.

**2.3.2. Antero-Posterior and Supero-Inferior Landmark Identification**

Using the central slab identified above, the remaining two coordinates are determined.

1. **Mean Coronal Image:** A representative mean coronal image, **I**_cor_, is generated by averaging all the sagittal slices within a small region around the midsagittal slice. This reduces noise and provides a clear view of the central anatomy.

**I**_cor_ = (1 / (**c**₂_max_ - **c**₂_min_ + 1)) × Σ **CT**(:, s, :) for s = **c**₂_min_ to **c**₂_max_

2. **Anatomical Midline (x-coordinate):** The patient's anatomical midline (c₁) is identified within a posterior region of this coronal image. This is achieved by calculating the intensity-weighted centroid along the horizontal axis, which effectively localizes the low-intensity signal of the gluteal cleft.

c₁ = C_x_(**I**_cor_(:, posterior region))

3. **Superior-Inferior Position (z-coordinate):** Finally, the z-coordinate (c₃) is determined. A Maximum Intensity Projection (MIP) coronal image is created. A 1D vertical intensity profile is extracted from this MIP, centered at the midline position c₁. The z-coordinate, c₃, is set as the most superior position in this profile where the intensity exceeds a threshold of 150 HU (indicating dense bone), minus a fixed offset of 20 voxels (an approximate estimation of the prostate location from the gluteal cleft).

**2.3.3. Initial Estimate Determination**

The three voxel coordinates (c₁, c₂, c₃) are combined and transformed from the image coordinate system into the physical, real-world coordinate system using the image's affine transformation matrix. This results in the final 3D estimate, **x̂**₀, which is used to initialize the main optimization grid search.

#### 1.2.4 Multi-Modal Cost Function Optimization

The final alignment was achieved through optimization of a composite cost function that balanced geometric accuracy with statistical intensity correspondence using **x̂**₀.

**Joint Probability Distribution:** A critical component of the optimization was a pre-computed joint probability density function P(I_CT_, I_T2_) that statistically modeled expected intensity relationships between CT and T2 voxel values, learned from the entire atlas cohort and applied within a 5mm radius of the prostate (V_ROI_),.

**Composite Cost Function:** The optimization minimized a weighted combination of intensity similarity and multi-modal correspondence terms:

Cost(x̂) = w_MSE_ ⋅ C_MSE_(x̂) + (1 - w_MSE_) ⋅ C_PDF_(x̂)

where x̂ represents the 3D translation vector being optimized (**x̂**₀).

**Intensity Similarity Term:** The Mean Square Error (MSE) component measured normalized intensity differences between the patient's transformed CT and the atlas CT:

C_MSE_(x̂) = (1 / |V_ROI_|) Σ [( I_CT_,_patient_(v + x̂) - A_CT_(v) ) / σ_CT_(v) ]²

This term ensured geometric alignment by comparing standardized intensity patterns within the region of interest.

**Multi-Modal Correspondence Term:** The PDF component enforced statistical consistency between co-located CT and T2 intensities:

C_PDF_(x̂) = - Σ log P(I_CT,patient_(v + x̂), I_T2,patient_(v))

This term leveraged the learned statistical relationships to constrain the registration using information from both imaging modalities simultaneously.

**Two-Pass Optimization Strategy:**

Pass 1 (Coarse Grid Search): A wide-range grid search prioritizing geometric alignment using only MSE (w_MSE_ = 1.0) within a large radius ROI around the prostate to robustly identify the general alignment region in each direction:

Δx ∈ {-30, -20,…,30} mm

Δy ∈ {-30, -20,…,30} mm

Δz ∈ {-60, -50,…,60} mm

This coarse alignment establishes an initial transformation that brings the patient’s CT into approximate alignment with the atlas prostate centroid.

Pass 2 (Fine Gradient-Based Search): A refined gradient-based optimization initiated from the best Pass 1 position. This stage balanced both cost terms (w_MSE_ = 0.5) within a tightened 5mm radius ROI focused on the prostate region, achieving precise final alignment using Nelder-Mead simplex optimization.

#### 1.2.5 Final Transformation and Registration Output

The optimization process yielded the final translation vector (T_reg_), defining the optimized translation matrix T_CT-trans_. The complete transformation mapping the patient's native CT space to atlas space combined the initial scaling with the optimized translation:

T_CT_→Atlas = T_CT-trans_ ⋅ T_scale_

**Primary Registration Output:** The algorithm's main output was the registration matrix T_reg_, which maps the patient's native T2 space to their native CT coordinate system:

T_reg_ = T_T2-initial_ ⋅ (T_CT_→Atlas)⁻¹

This final transformation matrix enables accurate resampling of the T2-weighted volume and prostate segmentation into the patient's planning CT coordinate system, facilitating precise multi-modal treatment planning by leveraging the statistical relationships learned from the training atlas cohort.

Supplementary Table 1: Imaging Acquisition Parameters

| **MRI Parameters** | | | |  | |  | |  |  |
| --- | --- | --- | --- | --- | --- | --- | --- | --- | --- |
| **Parameters** | | **GE SIGNA Premier** | | **GE Signa HDxt** | | **SIEMENS Espree** | | **SIEMENS Magnetom Vida** | **SIEMENS Skyra** |
| Pulse sequence | | SE | | SE | | SE | | SE | SE |
| TR (ms) | | 3400 | | 3900 | | 4000 | | 7000 | – |
| TE (ms) | | 100 | | 100 | | 90 | | 120 | 100 |
| FOV | | 1024×1024 pixels | | 512×512 pixels | | 512×512 pixels | | 640×640 pixels | 320×320 pixels |
|  | | (160.05x160.05mm) | | (199.99x199.99mm) | | (180.00x180.00mm) | | (180.00x180.00mm) | (230.00x230.00mm) |
| Slice Thickness  (mm) | | 3 | | 3 | | 3 | | 3 | 2 |
| Field Strength  (T) | | 3 | | 3 | | 1.5 | | 3 | 3 |
|  | **CT Parameters** | | | |  | |  | | |
|  | **Parameters** | | **GE Discovery CT590 RT** | | **Philips Big Bore** | | **SIEMENS SOMATOM Confidence** | | |
|  | kVp | | 120 | | 140 | | 120 | | |
|  | Tube Current  (mA) | | 600 | | 100 | | 160 | | |
|  | Exposure Time  (s) | | 70 | | 120 | | 180 | | |
|  | FOV | | 512×512 pixels | | 512×512 pixels | | 512×512 pixels | | |
|  |  | | (500.00x500.00mm) | | (700.00x700.00mm) | | (550.00x550.00mm) | | |
|  | Slice Thickness  (mm) | | 1.25 | | 2 | | 2 | | |

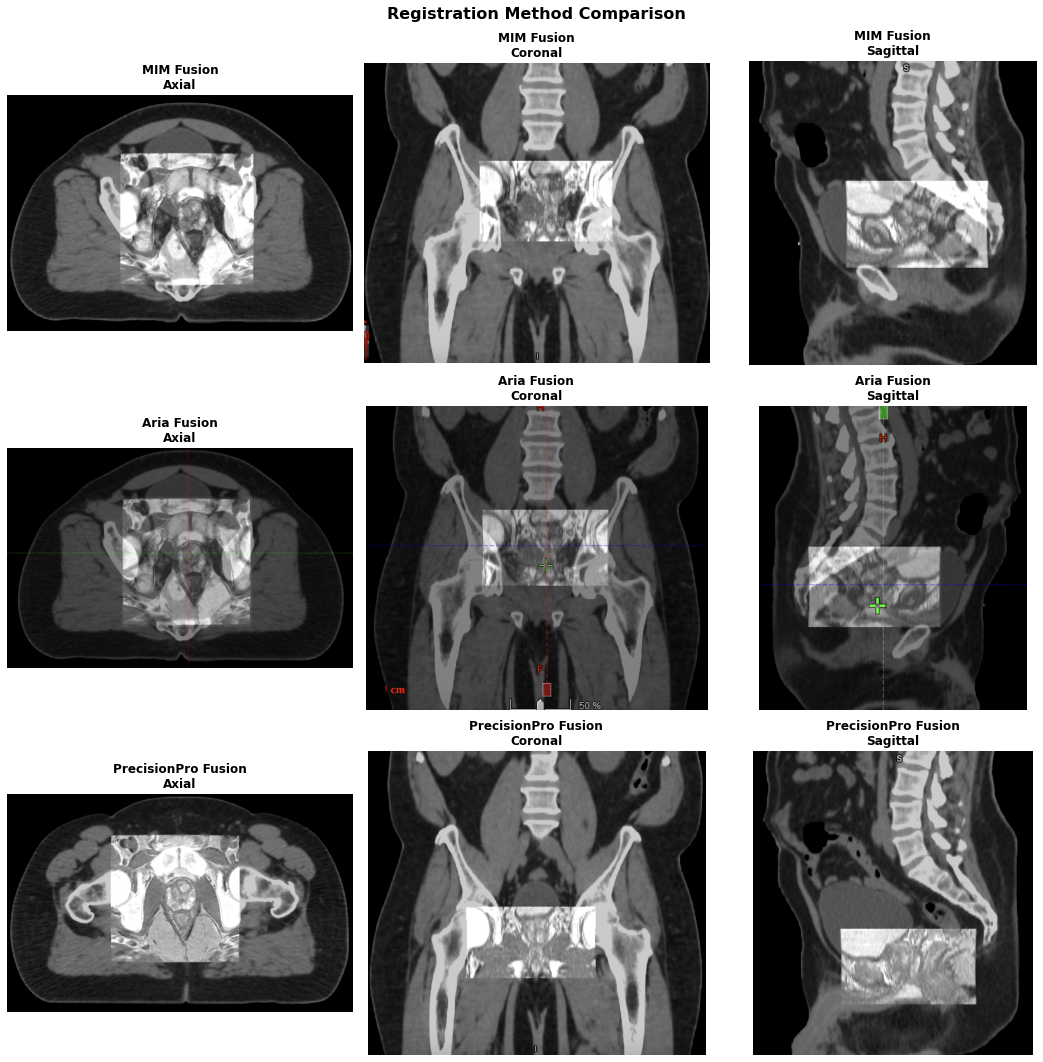

**Supplementary Figure 1: Comparison of MRI/CT fusion images using three registration methods across axial, coronal, and sagittal planes.** An example patient case comparing 3 different automated registration methodologies for MRI-CT registration. The top row shows results from MIM Fusion, the middle from Aria Fusion, and the bottom from PrecisionPro Fusion. Each column corresponds to a different anatomical view: axial, coronal, and sagittal. PrecisionPro Fusion displays strong alignment of both soft tissue and bony structures, while MIM and Aria Fusion both struggle to accurately localize the prostate. PrecisionPro Fusion displays the most precise anatomical correlation for prostate radiotherapy treatment planning.

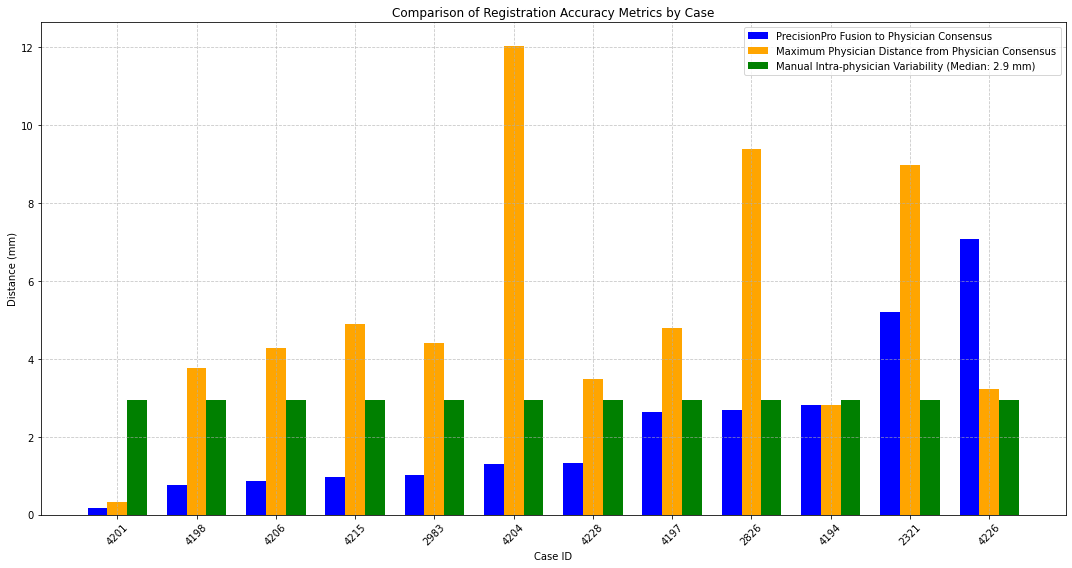

**Supplementary Figure 2: Comparative analysis of PrecisionPro Fusion’s registration accuracy versus manual physician registration variability.** Boxplot illustrating the performance of the PrecisionPro Fusion algorithm (blue) from the average of physicians’ registration matrices (consensus) compared to maximum physician deviations from the physician consensus (yellow) and manual intra-physician consistency (green). In 10 out of 12 patient cases (83%), distance of PrecisionPro Fusion’s registration to the physician consensus was smaller than intra-physician variability for manual registration of that case, indicating clinical reliability comparable to expert human performance.

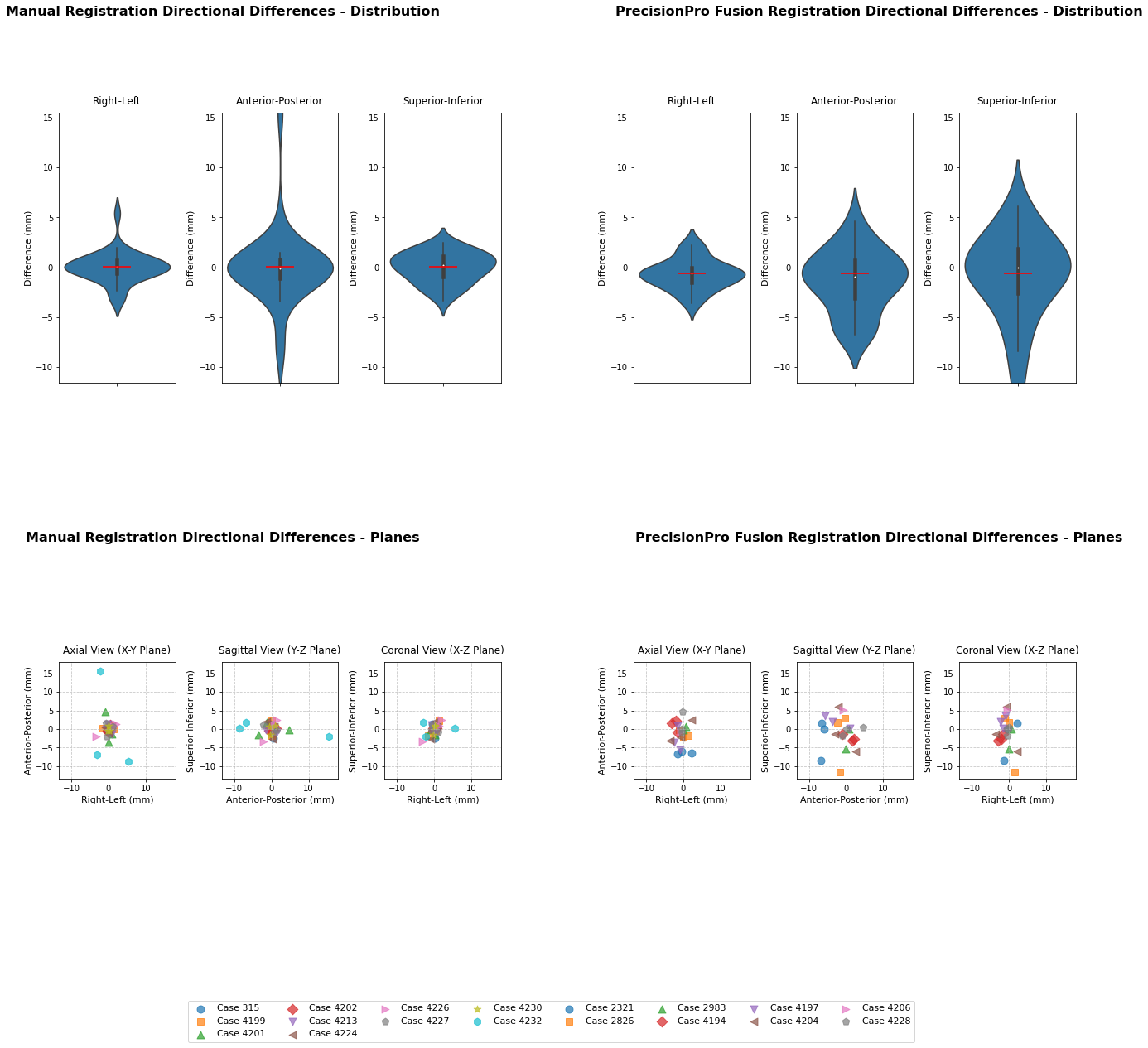

**Supplementary Figure 3: Multi-dimensional directional analysis of registration variability between physicians and PrecisionPro Fusion.** Four-panel visualization depicting: (Top left) Spatial distribution of inter-observer directional variability from the average of physicians’ registration matrices (consensus); (Top right) Directional variability in manual adjustments to PrecisionPro Fusion registrations; (Bottom left) Scatter plot depicting physician directional variability from the physician consensus across the patient cohort; (Bottom right) Case-by-case directional variability of PrecisonPro Fusion manual adjustments. Manual inter-observer registration directional variability from the centroid showed minimal systematic bias with median displacements of 0.1 mm (IQR: -0.6, 0.7 mm) in the right-left direction, -0.1 mm (IQR: -1.1, 0.8 mm) in the anterior-posterior direction, and 0.3 mm (IQR: -0.9, 1.1 mm) in the superior-inferior direction.

PrecisionPro Fusion adjustment directional variability demonstrated slightly larger displacements with medians of -0.6 mm (IQR: -1.5, 0.0 mm) in the right-left direction, -0.9 mm (IQR: -3.1, 0.7 mm) in the anterior-posterior direction, and 0.0 mm (IQR: -2.6, 1.9 mm) in the superior-inferior direction. Quantitative assessment demonstrates systematic bias across all three anatomical planes (right-left, anterior-posterior, and superior-inferior) for both manual and PrecisionPro Fusion adjustments, with more noticeable variability in physician corrections to PrecisionPro Fusion. Notably, there is a noticeably higher adjustment magnitude to PrecisionPro Fusion registrations specifically in the superior-inferior direction, suggesting registration challenges within this anatomical plane.
